# An Empirical Investigation into Measurement and Determinants of Healthcare Access in Rural Nigeria: A Multidimensional Perspective

**DOI:** 10.64898/2026.02.13.26346240

**Authors:** A. M Yaqoob, K.K Salman

## Abstract

Despite numerous reforms and initiatives within the healthcare system, Nigeria’s performance on universal health coverage (UHC) indicators remains sub-optimal. This study aims to empirically examines multidimensional access to healthcare and identifies the key determinants influencing levels of access among rural households in Nigeria. We utilize cross-sectional data from 625 rural households collected between June and October 2022. Multidimensional access to healthcare was measured using a 10-item instrument capturing participants’ psychological experiences of access. Data were analyzed using Confirmatory Factor Analysis (CFA) and an Instrumental Variable ordered probit (IV-Oprobit) model.Two items were excluded from the scale due to non-convergence and high residual correlations. The resulting eight-item scale demonstrated acceptable internal consistency (Cronbach’s α = 0.70). Among the four dimensions of access, availability recorded the lowest mean score, while geographical accessibility had the highest. Overall, only one in eight rural households achieved adequate access to healthcare, with the majority either vulnerable to or experiencing inadequate access. Monthly per capita expenditure (MPCE), household size, educational attainment, region of residence, facility operating hours, perceived quality of care, and waiting time are all significantly associated with the probability of reporting adequate, moderate, or inadequate access to healthcare. While expanding financial protection through broader health insurance coverage for rural populations remains critical, such efforts must be complemented by interventions that address persistent supply-side constraints to healthcare access.

**Key messages:**

- At 38, Nigeria has one of the lowest Universal Healthcare Coverage (UHC) index in sub-Saharan Africa suggesting under-performance in healthcare system.
- An important but under-explored challenge in the pursuit of UHC particularly in LMIC countries is the conceptualization and measurement of healthcare access.
- Findings show that, overall access to healthcare remains inadequate with one out of eights households in rural Nigeria achieved adequate access.
- The strong influence of consumption expenditure underscores the continued dominance of out-of-pocket (OOP) financing, emphasizing the need to expand financial protection through health insurance coverage.
- Distinguishing nominal from quality-adjusted access and embedding multidimensional access measures into routine health surveys can strengthen evidence-based planning and support targeted efforts to reduce rural health inequities.

## Introduction

Despite global progress toward Universal Health Coverage (UHC), gains in sub-Saharan Africa have lagged considerably. While the global UHC service coverage index increased from 54 in 2000 to 71 in 2021, reflecting improvements across key service areas, the corresponding estimate for sub-Saharan Africa reached only 43 over the same period (WHO & World Bank, 2023). As a result, fewer than half of the region’s population has access to essential health services (Porgo *et. al*., 2024). These regional aggregates, however, mask substantial cross-country variation, underscoring the importance of country-level analysis for identifying health system gaps and informing policy responses.

Within sub-Saharan Africa, Nigeria continues to underperform on UHC indicators. In 2021, Nigeria’s UHC service coverage index stood at 38—well below the regional average and lagging behind several peer countries, including Ghana, Senegal, and Côte d’Ivoire (WHO, 2025; Population Reference Bureau, 2025). This low level of coverage implies that a large share of the population continues to face barriers in accessing essential health services. These challenges are particularly pronounced in rural areas, where limited health financing, workforce shortages, socioeconomic deprivation, and uneven service distribution constrain access to quality care (Nwankwo *et. al*., 2022).

Empirical evidence consistently shows stark rural–urban disparities in healthcare access in Nigeria. Coverage of antenatal care, skilled birth attendance, and routine immunization is substantially lower among rural populations compared with urban residents (Adewuyi & Zhao, 2020). These gaps contribute to preventable morbidity and mortality and reflect broader inequalities in health outcomes (Onakalu *et. al*., 2025). Despite multiple policy initiatives—including social health insurance schemes, immunization programs, and performance-based financing—the overall performance of Nigeria’s health system remains weak. Life expectancy remains below regional and global averages, and Nigeria accounts for more than one-quarter of global maternal deaths (WHO *et. al*., 2023; Weitensfielder et al., 2024). These outcomes reflect a combination of demand- and supply-side barriers, including poverty, distance to facilities, workforce shortages, poor service quality, and uneven facility distribution.

An important but underexplored challenge in the pursuit of UHC—particularly in low- and middle-income countries—is the conceptualization and measurement of healthcare access itself (Ensor and Cooper, 2004; Kruk *et. al*., 2018). Although healthcare utilization is related to access, it is often used inappropriately as a proxy, despite clear conceptual distinctions. Utilization alone cannot capture the multidimensional nature of access or the barriers that prevent individuals from obtaining needed care (Whitehead *et. al*., 1997). Healthcare access is inherently multidimensional, encompassing availability of services, geographic accessibility, financial affordability, service acceptability, and quality of care (Levesque *et. al*., 2013). Geographic accessibility reflects the spatial relationship between service users and providers, considering distance, travel time, transportation, and cost (Tanou *et. al*., 2021). Financial accessibility relates to individuals’ ability to pay for services without incurring financial hardship—a core principle of UHC (WHO, 2025). Acceptability captures the extent to which health services align with users’ social and cultural expectations, with poor provider–user relationships often constituting a major barrier in low-resource settings (Palmer, 2007).

Despite growing interest in healthcare access, relatively few studies adopt a truly multidimensional measurement approach (Musau *et. al*., 2025). Much of the empirical literature relies on single indicators—such as service utilization, insurance coverage, or out-of-pocket spending—which provide fragmented and potentially misleading representations of access (Edeh, 2022; Okunogbe *et. al*., 2022; Onah *et. al*., 2023). While these studies identify important socioeconomic gradients, they fail to capture how multiple access barriers interact simultaneously within households. Moreover, existing research in Nigeria and similar contexts remains largely sector-specific, focusing predominantly on maternal and child health services rather than access across the broader continuum of care (Afulani *et. al*., 2023; Adam *et. al*., 2023).

This study addresses these gaps by developing and applying a valid and reliable psychometric-based measure of healthcare access that captures overlapping dimensions of access and their distribution within rural populations. Using primary data from rural Nigeria, the study empirically examines multidimensional access to healthcare and identifies key determinants shaping different levels of access. By moving beyond utilization-based measures, this approach provides more precise evidence to inform targeted interventions and support equitable progress toward universal health coverage.

## 2. Materials and Methods

### 2.1 Study Settings

The survey was conducted in rural Nigeria between May and October 2022. Rural Nigeria is home to approximately 50–60% of the country’s population, characterized by low population density, dispersed settlements, and limited infrastructure (World Bank, 2024). Most households rely on subsistence agriculture and face lower income and educational attainment compared with urban areas, which constrains access to healthcare. Health facilities are sparse, often under-resourced, and located far from many communities, resulting in geographic and service-quality barriers to care (NPC & ICF, 2023). These demographic, socioeconomic, and health system characteristics make rural Nigeria a relevant setting for studying multidimensional healthcare access. The average rural population share over the post-independence period is 64.55%, and this historical mean provides the basis for our sample size estimation (Macrotrend, 2025).

### 2.2 Sample size and design

The sample size for this study was determined using the International Fund for Agricultural Development (IFAD) protocol (Equation 1). Based on the formula, a 98% confidence level (Z = 2.33), a margin of error of 4.5% (0.045), and an estimated proportion of rural households of 65% (0.65) yielded a calculated sample size of 610 households. Adjustments for design effects and a contingency allowance increased the final sample size to 624 households. Of these, 505 completed questionnaires (approximately 81% response rate) were valid and included in the analysis; the remainder were excluded due to missing or inconsistent information. To address potential non-response bias, inverse-probability weighting was applied. Response propensities were estimated via logistic regression, and the resulting weights were combined with the design sampling weights. To prevent extreme values from disproportionately influencing results, weights exceeding the 99th percentile were trimmed. Following IFAD’s methodological standards, a 98% confidence level and a 4.5% margin of error were used to enhance statistical rigor, ensuring narrower uncertainty intervals, and improves the reliability of estimates, thereby supporting evidence-based decision-making.

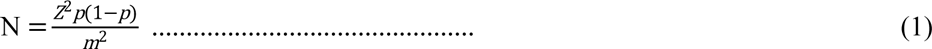

where N = the sample size, other parameters are as previously defined.

A multistage sampling procedure was employed to select 624 rural households. In the first stage, three of Nigeria’s six geopolitical zones—Southeast, Southwest, and Northeast—were randomly chosen. In the second stage, two states were randomly selected from each chosen zone: Imo and Ebonyi (Southeast), Oyo and Ekiti (Southwest), and Taraba and Borno (Northeast). In the third stage, two rural local government areas (LGAs) were randomly selected from each state. The fourth stage involved randomly selecting two wards from each LGA, yielding a total of 24 wards. Finally, 26 households were randomly chosen from each ward, resulting in a total sample of 624 households.

### 2.3 Data Collection

Primary data were collected using a semi-structured questionnaire administered through oral interviews. Enumerators were carefully recruited and trained to ensure consistent administration of the instrument. Before data collection, participants were informed of the study objectives and assured that their responses would be treated with strict confidentiality and anonymity. They were also informed of their right to withdraw from the study at any time, and verbal informed consent was obtained from those willing to participate. Responses were collected from the household head or another household member aged 18 or above who was knowledgeable about the household. The questionnaire comprised two main sections. The first section captured household demographic and socioeconomic characteristics, including age, gender, household size, marital status, education, primary occupation, income source, total household expenditure, and wealth indicators. The second section focused on health and healthcare, covering providers visited, type of illness experienced, out-of-pocket health expenditure, average waiting time, perceived quality of healthcare, facility operating hours, and household’s subjective evaluation of healthcare accessibility across ten items.

### 2.4 Measurement Instrument

The healthcare access questionnaire was developed in collaboration with public health research experts. Face and content validity were assessed prior to data collection. Face validity was evaluated among 30 conveniently selected members of the target population, who provided feedback on item clarity, wording, and relevance. Based on their input, ambiguous items were revised to improve comprehension. Content validity was assessed using both qualitative and quantitative approaches. Qualitatively, a multidisciplinary panel of ten experts—comprising four maternal and child health clinicians/researchers, two health systems specialists, one measurement/biostatistics expert, two programme implementers, and one community representative—reviewed the questionnaire. Their feedback informed item refinement, removal of redundancy, and ensured adequate coverage of all access dimensions. Quantitatively, the Content Validity Ratio (CVR) and Content Validity Index (CVI) were computed. Items with CVR ≥ 0.62 and CVI ≥ 0.78 were retained, while items below these thresholds were revised.

Construct validity was examined using Confirmatory Factor Analysis (CFA) to assess whether items loaded appropriately onto the hypothesised access sub-dimensions. Factor loadings ≥ 0.70 indicated strong convergent validity, while loadings ≥ 0.50 were considered acceptable (Kline, 2016; Hair *et. al*., 2019). Model fit was evaluated using established fit indices within CFA and Structural Equation Modelling frameworks (Hu & Bentler, 1999). Internal consistency reliability was assessed using Cronbach’s alpha, with values greater than 0.60 considered acceptable (Daud *et. al*., 2018).

The access questionnaire consisted of ten items adapted from a previously non-validated survey (Olatunji *et. al*., 2013). Items were organized into four access domains: geographical accessibility (one item), physical availability (four items), financial affordability (two items), and acceptability (three items), consistent with established access frameworks (Penchansky & Thomas, 1981). Quality of care was not treated as a separate domain, as it is integral to all access dimensions and reflects the technical capacity of health services to influence health outcomes (Peters *et. al*., 2008).

Respondents identified the three most important healthcare providers visited in the three months preceding the survey. Although longer recall periods have been used in similar studies (Hosein-Estidarjani *et al*., 2021), a three-month recall period was chosen to minimize recall bias, particularly in settings with low literacy and limited record-keeping. Participants rated their access to each provider using a five-point Likert scale ranging from strongly disagree (1) to strongly agree (5). A composite indicator of healthcare access was calculated by averaging responses across the ten items for each provider, yielding scores ranging from 10 to 50.

### 2.5 Definition of variables

#### 2.5.1 Dependent variable

The outcome variable was the healthcare accessibility score, derived from a set of multidimensional access indicators. This continuous measure was calculated as the average of non-missing item responses, each scored on a five-point scale (1–5), for both the individual latent access dimension and the overall access construct. Using midpoint-based cut-off values, respondents were classified into three ordered categories: inadequate access (mean score < 3.0), moderate access (3.0 ≤ mean score < 4.0), and adequate access (mean score ≥ 4.0).

#### 2.5.2 Independent variables

The selection of independent variables was guided by the Andersen health behaviour model and evidence from prior empirical studies (Andersen, 1995; Pengid *et. al*., 2021; Mosha *et. al*., 2025). They include predisposing, enabling and need variables. The **Predisposing variables**: marital status, household size, educational attainment and primary occupation. **Individual or household level Enabling variables:** income using per capita expenditure as proxy. Per capita expenditure rather than income was used because it is often considered a better proxy than income since it is less sensitive to short-term fluctuations (Wossen *et. al*., 2019). The enabling variables that are related to the health system and external environment include: average waiting time, facility average operating hours, cost of care, perceived quality of health services, distance to the nearest health facility and geographical region (contextual variation). The **need variables** include: sex, age, and health condition. The descriptive statistics of the variables are presented in Table 1.

**Table 1:** Summary statistics of variables.

| **Variable** | **Mean** |
| --- | --- |
| Sex of household head (1= male, 0 otherwise) | 0.89 |
| Age of household head (years) | 50.18 (11.74) |
| Marital status (1= Married, 0 otherwise) | 0.89 |
| Household Size (#) | 7.62 (3.57) |
| Educational attainment (1= primary & below, 2= secondary, 3= tertiary) | 1.88 |
| Primary occupation (1= farming, 0 otherwise) | 0.49 |
| Per capita monthly expenditure (₦) | 16,241.21(12,945.4) |
| Distance to the nearest health facility (km) | 4.89 (6.95) |
| Average waiting time (hours) | 0.64 (0.57) |
| Average operating hours (hours) | 19.29 (3.89) |
| Average rating of quality of health service<br>(1= fair, 0 otherwise) | 0.65 |
| Healthcare accessibility score (#) | 3.02 (0.47) |
| Mean community-level asset | 557.50 (1238.74) |
| Out-of-pocket (OOP) health expenditure (#) | 26,177.1 (33,640.45) |
| Health condition (1= chronic, 0 otherwise) | 0.79 |
| Region of residence (1=North, 0 otherwise) | 0.29 |

Values in parenthesis are standard deviation reported only for continuous variable.

The variable instrumenting per capita expenditure is mean community-level asset. The key role of community-level asset endowment in influencing the socioeconomic environment within which households operate and in creating local economic opportunities for people informs the choice of this variable. Previous studies have shown that such community wealth indicators are strongly correlated with individual or households welfare levels, since wealthier communities tend to have higher employment opportunities, better markets, and greater access to productive resources (Filmer and Pritchett, 2001; Montgomery *et. al*., 2000). With the exception of assets that are used for transportation including vehicle, motorcycle, bicycle, such community-level asset variable is assumed to have a direct influence on household’s level of income denoted by consumption expenditure without directly influencing accessibility to health service. The mean Community-level asset ---leaving own asset out ---was calculated using the formula below:

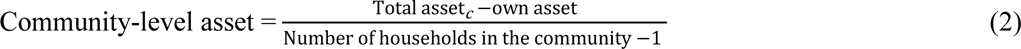

### 2.6 Data Analysis

Descriptive statistics were used to summarize key characteristics of the study population. Confirmatory factor analysis within a structural equation modelling framework was applied to validate the four-factor psychometric measure of multidimensional access to healthcare. Determinants of healthcare access were examined using instrumental variable (IV) models to account for potential endogeneity. Access was analyzed both as an ordered (inadequate, moderate, adequate) and as a continuous outcomes, estimated using IV ordered probit and linear IV (two-stage least squares) specifications, respectively. Monthly per capita household expenditure (MPCE) was treated as endogenous, reflecting the likely joint determination of household welfare and healthcare access. Mean community-level asset ownership was used as an instrument for MPCE, capturing exogenous variation in local economic conditions and assumed to affect healthcare access only through MPCE. For the ordered outcome, access and consumption expenditure equations were estimated jointly using full-information maximum likelihood via the Conditional Mixed Process estimator. The significance of the reported coefficient of antrho statistics is a confirmation of the suspected endogeneity. For the linear specification, endogeneity and instrument strength were examined using standard Durbin–Wu–Hausman and weak-identification tests. All analyses were conducted using Stata version 17.

## 3. Results

### 3.1 Internal Validity of the Measurement Instrument

The result of the Fit Indices for Confirmatory Factor Analysis was presented in Table 2. The fitness of the model as indicated by TLI (0.96); CFI (0.97); SRMR (0.05) RMSEA (0.05) and X2/df (2.42), suggests that they are all within good or acceptable regions confirming that the model adequately fits the data. Further, the standardized factor loadings for the final 8 items measuring the latent 4 sub-constructs -Availability, Geographical Accessibility, Affordability and Acceptability are in the range of 0.54 to 0.84 (see the appendix) implying that each of the measurement items capture sufficient variation in their respective latent sub-constructs.

**Table 2:** Fit Indices for Confirmatory Factor Analysis model.

| Model fit Indices | X<sup>2</sup>/df | RMSEA | SRMR | CFI | TLI |
| --- | --- | --- | --- | --- | --- |
| Confirmatory Factor Analysis | 2.42 | 0.05 | 0.05 | 0.97 | 0.96 |
| Threshold for acceptable fit | < 5 | < 0.08 | < 0.08 | ≥ 0.90 | ≥ 0.9 |
| Threshold for good fit | < 2 | ≤ 0.05 | ≤ 0.05 | ≥ 0.95 | ≥ 0.95 |

### 3.2 Internal Consistency of the Measurement Instrument

The internal consistency of the measurement instrument was presented in Table 3. The Cronbach’s alpha for “Availability” “Affordability/Accessibility” and “Acceptability” dimensions were 0.84, 0.61 and 0.64 respectively indicating that the items are reliable indicators. The Cronbach’s alpha for “Geographical Accessibility” dimension could not be assessed for a single-factor single indicator because Cronbatch’s alpha requires multiple indicators to estimate inter-item consistency. Nonetheless, the Cronbach’s alpha for the overall access was 0.7 indicating an acceptable level of reliability (Martin and Emily, 2013). Furthermore, the ICC value obtained for the questionnaire was 0.80 indicating an acceptable level of stability.

**Table 3:** The Cronbach’s alpha coefficient for Measurement Instrument.

| Access Dimensions | Number of items | Cronbach's alpha coefficient |
| --- | --- | --- |
| Availability | 3 | 0.84 |
| Affordability | 2 | 0.61 |
| Geographical accessibility | 1 | - |
| Acceptability | 2 | 0.64 |
| Overall access | 8 | 0.70 |

### 3.3 Evaluating multidimensional access to healthcare in rural Nigeria

The participants’ mean scores across the four evaluated dimensions of healthcare access ranged from 2.61 to 3.66 (Table 4). Among these dimensions, Availability recorded the lowest mean score (2.61 ± 0.73), followed by Acceptability (2.93 ± 0.62). In contrast, Geographical accessibility had the highest mean score (3.66 ± 0.75), indicating comparatively better performance on this dimension. The low mean score for the availability dimension reflects deficiencies in the physical presence of health facilities, as well as inadequacies in essential medical equipment, supplies, and healthcare personnel. Similarly, the acceptability dimension—which captures the extent to which health services align with users’ social and cultural expectations—was also rated as insufficient. Conversely, the relatively higher mean score for geographical accessibility suggests a moderate level of access, which may be partly explained by the presence of informal healthcare providers that serve as alternative sources of care in rural communities. Overall, the composite mean score for psychometric-based access to healthcare was 3.02 ± 0.47. Using the scale midpoint as a benchmark, only 11.9% of rural residents were found to have adequate access to quality healthcare across all dimensions—availability, geographical accessibility, affordability, and acceptability—while the majority were classified as being at risk of inadequate access, either due to vulnerability or clear insufficiency.

**Table 4:**
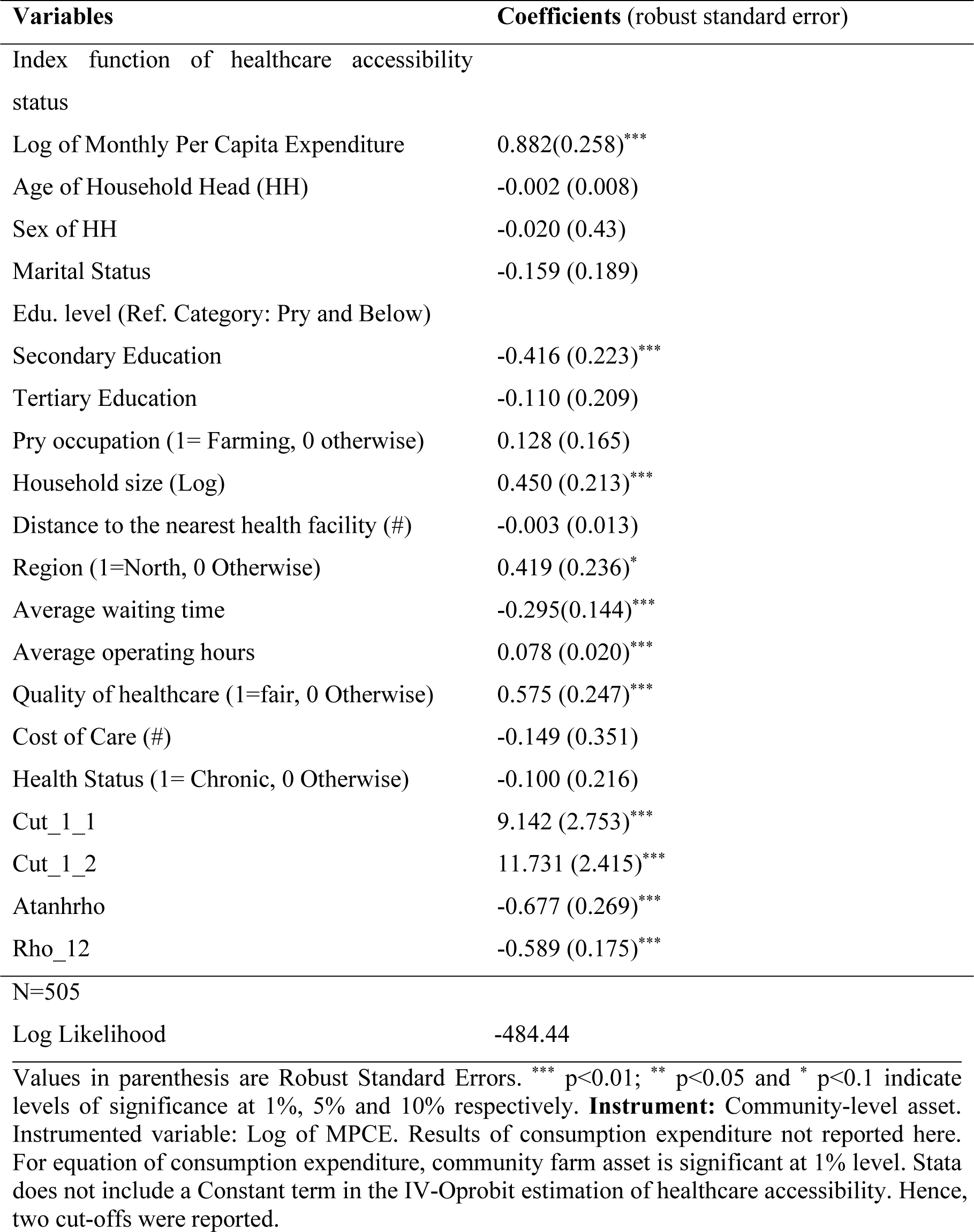
Results of IV-OProbit Model (Conditional Mixed Process-CMP)

### 3.4 Factors determining level of multidimensional access to healthcare

Table 4 presents the estimated result of IV-ordered probit. As reported in the table, the coefficient of atanhrho, which measures the correlation between the error terms of structural and reduced-form equation is significant [β = -0.677; p < 0.01]. This indicates a strong evidence of endogeneity in our underlying model, justifying the use of a full simultaneous equation structure. Out of seven coefficients of the significant variables, the direction of effects for the five variables is positive. They include log of monthly per capita expenditure (MPCE), household size, region of residence, average operating hours of the facility and average rating of the quality of healthcare. This implies increasing effects on the probability of reporting highest category (adequate access) and decreasing effect on the probability of reporting lowest category (inadequate access). For the remaining two variables --secondary education and average waiting time, the direction of effects is negative implying decreasing effect on the probability of reporting highest category (adequate access) and increasing effects on the probability of reporting lowest category (inadequate access). For the middle category [P(Y=1)], the sign of the coefficient is ambiguous, normally, one would expect a sign change in the intermediate category based on the single crossing effect. In the absence of formal test for validity of the instrument in non-linear model of this type, the significance of the instrument (z) indicates that the instrument is valid in the present study (Greene, 2012).

The instrumental variable—mean community-level asset—is negative and statistically significant. This indicates that individuals living in relatively wealthier communities are, on average, less likely to utilize public health services. Although this finding may seem counter-intuitive, it is consistent with evidence from the health-seeking behavior literature, which emphasizes the nuanced influence of community-level economic contexts on patterns of healthcare use (Kruk *et. al*., 2018). A plausible explanation is a substitution effect, whereby residents of economically better-off areas have greater access to alternative sources of care—such as private health facilities, pharmacies, or specialist services—and consequently depend less on public or informal healthcare providers. It is important to note that, within the framework of the ordered probit model employed in this study, the estimated coefficients indicate only the direction of the relationships and do not convey the magnitude of the effects. Consequently, marginal effects evaluated at the mean values of the explanatory variables are reported in Table 5 to provide a more informative interpretation of the results.

**Table 5:** Marginal Effects of IV-Oprobit Model.

| Variables | $\frac{\partial(Y=0)}{\partial x}$ | $\frac{\partial(Y=1)}{\partial x}$ | $\frac{\partial(Y=2)}{\partial x}$ |
| --- | --- | --- | --- |
| Log of Monthly Per Capita Expenditure (MPCE) | -0.159 (0.078)*** | -0.010 (0.021) | 0.169 (0.068)*** |
| Age of HH | 0.0003 (0.001) | 0.019 9 (0.012) | -0.0003 (0.001) |
| Sex of HH | 0.004 (0.074) | 0.0002 (0.004) | -0.004 (0.079) |
| Marital Status | 0.029 (0.035) | 0.002 (0.005) | -0.030 (0.037) |
| **Educational Level** |  |  |  |
| (Ref. Category): Primary |  |  |  |
| Education & below) | - | - | - |
| Secondary Education | 0.081 (0.044)** | -0.007 (0.014) | -0.074 (0.037)*** |
| Tertiary Education | 0.019 (0.037) | 0.003 (0.0006) | 0.022 (0.043) |
| Primary Occupation | -0.023 (0.030) | -0.001 (0.003) | 0.025 (0.031) |
| Log of Household Size | -0.081 (0.048)* | -0.005 (0.011) | 0.086 (0.047)* |
| Distance to the Nearest Health Facility | 0.0005 (0.002) | 0.035 (0.164) | -0.606 (0.025) |
| Region of Residence | -0.075 (0.052) | -0.005 (0.010) | 0.081 (0.050)* |
| Average Waiting Time | 0.053 (0.030)*** | 0.003 (0.007) | -0.056 (0.030)*** |
| Average Operating Hours | -0.014 (0.004)*** | -0.086 (0.205) | 0.015 (0.003)*** |
| Perceived Quality of Health Service | -0.104 (0.032)*** | -0.0006 (0.016) | 0.110 (0.040)*** |
| Cost of Care | 0.268 (0.658) | 0.165 (0.476) | -0.284 (0.686) |
| Health condition | 0.018 (0.037) | 0.001 (0.004) | -0.019 (0.040) |

Note: Values reported in parenthesis are standard errors; \*\*\*, \*\* and \* represent levels of significance at 1%, 5% and 10% respectively.

**Table 6:**
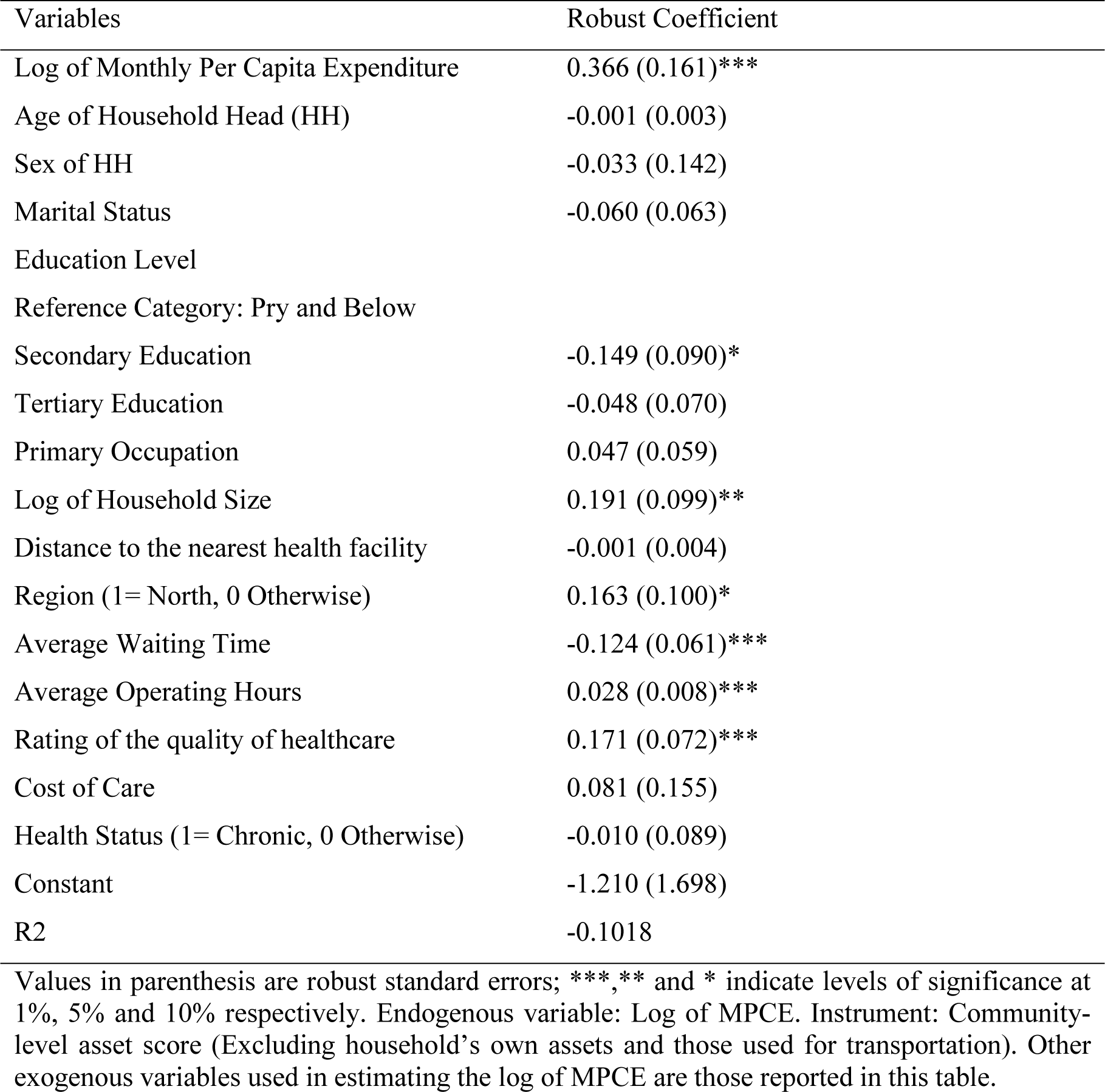
Results of Linear IV Regression of Log of MPCE on accessibility to healthcare.

| Variables | Robust Coefficient |
| --- | --- |
| Log of Monthly Per Capita Expenditure | 0.366 (0.161)*** |
| Age of Household Head (HH) | -0.001 (0.003) |
| Sex of HH | -0.033 (0.142) |
| Marital Status | -0.060 (0.063) |
| Education Level |  |
| Reference Category: Pry and Below |  |
| Secondary Education | -0.149 (0.090)* |
| Tertiary Education | -0.048 (0.070) |
| Primary Occupation | 0.047 (0.059) |
| Log of Household Size | 0.191 (0.099)** |
| Distance to the nearest health facility | -0.001 (0.004) |
| Region (1= North, 0 Otherwise) | 0.163 (0.100)* |
| Average Waiting Time | -0.124 (0.061)*** |
| Average Operating Hours | 0.028 (0.008)*** |
| Rating of the quality of healthcare | 0.171 (0.072)*** |
| Cost of Care | 0.081 (0.155) |
| Health Status (1= Chronic, 0 Otherwise) | -0.010 (0.089) |
| Constant | -1.210 (1.698) |
| R2 | -0.1018 |

Values in parenthesis are robust standard errors; \*\*\*,\*\* and \* indicate levels of significance at 1%, 5% and 10% respectively. Endogenous variable: Log of MPCE. Instrument: Community-level asset score (Excluding household's own assets and those used for transportation). Other exogenous variables used in estimating the log of MPCE are those reported in this table.

#### Individual/Household Characteristics

The marginal effect estimates of Monthly Per Capita Expenditure (MPCE) are significant [β= - 0.169; β= -0.159: p < 0.01] for upper and lower outcome categories respectively. This implies that for a rural household with an average MPCE of 16,241.2, a unit percentage point increase in consumption expenditure increases the probability of reporting adequate access in all forms--availability, accessibility, affordability and acceptability-- to healthcare by 16.9% and decrease the probability of reporting inadequate access by 15.9%, given other variables. For secondary education, the marginal effect estimates [β= -0.074; β = 0.081: p < 0.01] are significant for both upper and lower outcome categories respectively.

This implies that the probability of reporting adequate access to healthcare was 7.4% lower for respondents who completed secondary education relative to primary education, while the probability of reporting inadequate access was 8.1% higher. Regarding household size [β=-0.086; β = 0.081: p < 0.1], for a household with average size of 7, a member increase in size of the household leads to an increase in the probability of reporting adequate access to healthcare by 8.6 percentage point and a decrease in the probability of reporting inadequate access by 8.1 percentage point.

#### Healthcare-specific characteristics

The marginal effect estimate for waiting time at health facility are [β= -0.056; β= 0.053: p < 0.01] for upper and lower outcome categories respectively and significant. The implication is that, for an average waiting time of about 40 minutes, a unit percentage point increase in facility waiting time decreases the probability of reporting adequate access to healthcare by 5.6% and increases the probability of reporting inadequate access by 5.3%. Consistent with theoretical expectation, for an average 19 hours of operation of health facility, [β= -0.015; β= 0.014: p< 0.01], a unit percentage point increase in operating hours of the facility increases the probability of reporting adequate access to healthcare by 1.5% and reduces the probability of reporting inadequate access by 1.4%. In line with *a priori* expectation, the marginal effect estimates for quality of health service are [β= 0.110; β= -0.104: p< 0.1] and significant. This implies that, given other control variables, respondents who perceived good quality of healthcare--relative to those who perceived poor quality-- were 11% more likely to report adequate access to healthcare and 10.4% less likely to report inadequate access.

#### Contextual factor

Contrary to the expectation, the marginal effect estimates for the region of residence are [β= 0.081; β= -0.075: p< 0.01] and significant. This implies that, holding all other variables constant, being resident in rural northern Nigeria increases the probability of reporting adequate access to healthcare by 8.1 percentage points and reduces the probability of reporting inadequate access by 7.5 percentage points.

The marginal effects for the remaining variables are not statistically significant. While several covariates exhibit relatively large effect sizes, others display negligible and insignificant effects. In the case of households classified as having moderate access, the results appear to reflect the inherent ambiguity of the intermediate category, whose characteristics are not clearly differentiated by observed socioeconomic or demographic factors (Knudsen & Berg, 2024). As expected in nonlinear models of this nature, marginal effects for the middle outcome category are often insignificant, since changes in probabilities tend to be concentrated at the lower and upper tails of the distribution rather than in the intermediate range (Wooldridge, 2015).

### 3.5 Results of Instrumental Variable (IV) Linear Model

Qualitatively, the result of linear IV (2SLS) model are similar to that of full information maximum likelihood estimation of IV-Ordered probit model presented in the previous section (Table 5). The coefficient of log of MPCE is positive and significant. This implies that, with an increase in per capita household expenditure-a proxy for income-- in all forms, access to healthcare also increases indicating an improvement in accessibility status. Secondary education, household size (log), region of residence, average waiting time, average operation hours, perceived quality of healthcare are significant in both linear IV and IV ordered probit model. The direction of effect of variables are also similar to that reported in the previous model. In this case, the coefficients are also the marginal effects of the covariates providing more information regarding the size and direction of effects.

We conduct diagnostic tests to assess the strength and validity of the instrument as well as parameter identification. The null hypothesis of exogeneity was rejected (p < 0.01) confirming that endogeneity is a genuine concern in the present case. The Cragg-Donal (F= 22.58) test of validity and strength of the instrument, which exceeds the 10% maximal IV relative bias critical value of 16.38 indicates that the instrument is indeed valid. The Hansen J test of overidentification also indicates that the model is just identified which is expected given that only one excluded instrument was used. Further tests regarding the explanatory strength and relevance of the instruments indicates that our resulting IV estimates are reliable (see the appendix).

## 4. Discussion

This study developed and validated a multidimensional scale for measuring healthcare access in rural Nigeria and identified key predisposing and enabling-related determinants of access using instrumental variable approaches. The findings provide both a psychometrically robust measurement tool and policy-relevant insights into barriers to healthcare access in resource-constrained settings.

Content and face validity assessments led to the refinement of the original instrument, resulting in an eight-item scale judged by experts and respondents to be clear, relevant, and appropriate for measuring healthcare access. The use of both CVR and CVI criteria strengthened confidence in item retention decisions and ensured alignment with the underlying conceptual framework of access. These refinements are particularly important in rural and low-income contexts, where poorly specified indicators may fail to capture individual or household’s lived experiences of healthcare access.

Construct validity results from confirmatory factor analysis support a multidimensional structure of healthcare access, with acceptable model fit. While availability and acceptability emerged as distinct dimensions, the geographical accessibility item cross-loaded with affordability, necessitating the combination of these domains into a single latent construct. This overlap likely reflects the contextual reality of rural Nigeria, where financial capacity mitigates physical distance and transport barriers. Similar interactions between geographic and financial access have been documented in other low- and middle-income settings, underscoring the need for context-sensitive measurement of access. Future surveys could enhance construct separation by incorporating objective measures such as travel time or distance to health facilities.

The standardized factor loadings indicate strong associations for availability and moderate but significant associations for affordability, accessibility and acceptability, providing evidence of convergent validity. Reliability analysis further demonstrates acceptable internal consistency of the overall scale, alongside good stability over time as indicated by the intraclass correlation coefficient. Together, these results suggest that the instrument is suitable for population-based surveys and policy-oriented analyses of healthcare access.

From a policy perspective, the validated scale offers a practical tool for monitoring inequities in healthcare access and evaluating the effectiveness of interventions aimed at improving rural health service delivery. The observed convergence of financial and geographic barriers highlights the importance of integrated policies that address both affordability and physical access—such as transport subsidies, strategic facility placement, and financial protection mechanisms—rather than treating these constraints in isolation. Embedding such multidimensional measures in national health surveys can strengthen evidence-based planning and support progress toward universal health coverage in Nigeria and similar settings.

Household access to healthcare was assessed across four dimensions: availability, accessibility, affordability, and acceptability. Among these, accessibility recorded the highest mean score, indicating moderate access, whereas availability showed the lowest mean score, reflecting persistent deficiencies in the physical presence of health facilities, essential equipment, and skilled health personnel. This pattern is consistent with longstanding evidence of inadequate health infrastructure in rural Nigeria, where existing facilities often lack the basic resources required to deliver essential services (Oluwadare & Adepoju, 2023). These shortages continue to constrain the capacity of primary healthcare facilities to provide effective and timely care. The relatively higher score observed for geographical accessibility may reflect the widespread presence of informal healthcare providers in rural communities. In contexts where geographic distance and weak formal service provision limit access to public or private facilities, rural households frequently rely on informal and traditional providers. Such providers are often geographically proximate, socially embedded within communities, and offer flexible payment arrangements, thereby lowering practical barriers to care (Kumar *et. al*., 2022). Evidence from other low- and middle-income countries similarly shows that informal providers account for a substantial share of primary care in rural areas due to their affordability, convenience, and proximity (Onwujekwe *et al*., 2022). Despite moderate geographical accessibility, overall access to healthcare remains limited. Only one in eight rural households achieved adequate access across all four dimensions, while the majority experienced vulnerability or inadequate access. This finding aligns with previous studies documenting persistent geographic and socioeconomic inequalities in access to quality healthcare in Nigeria, particularly among rural populations (Okoli *et. al*., 2022). Together, these results highlight the gap between physical or perceived access to care and the availability of adequately resourced, high-quality health services in rural settings.

The IV-ordered probit results identify a set of economic, household, and service-related characteristics defined as the *Enabling* and *Predisposing* factors that significantly shape healthcare access among rural households in Nigeria. Monthly per capita expenditure (MPCE), household size, educational attainment, region of residence, facility operating hours, perceived quality of care, and waiting time are all strongly associated with the probability of reporting adequate, moderate, or inadequate access to healthcare.

Economic capacity emerges as a central determinant of healthcare access. Higher consumption expenditure—used as a proxy for household income—significantly increases the probability of reporting adequate access and reduces the likelihood of inadequate access. The negative sign observed for the intermediate category of moderate access is consistent with the single-crossing property of ordered response models, indicating a monotonic shift toward higher access as income increases. This finding aligns with theoretical expectations that consumption rises with income, thereby enhancing households’ ability to overcome financial, geographic, and service-related barriers to care. In Nigeria where healthcare financing is predominantly out-of-pocket (OOP) accounting for approximately 71% of total health expenditure, access to healthcare remains highly income-dependent (Mussa *et. al*., 2025). Similar evidence has been reported in recent studies examining healthcare utilization in low- and middle-income settings (Song *et. al*., 2025).

Educational attainment shows a more nuanced relationship with healthcare access. Relative to individuals with at most primary education, respondents with secondary education are less likely to report adequate access and more likely to report inadequate access. While this finding may appear counter intuitive, it likely reflects differences in expectations and decision-making regarding quality of care. Individuals with higher education may be more discerning in their choice of health facilities and less willing to utilize poorly resourced public primary healthcare centers that dominate rural areas. Consequently, they may forgo care or seek alternatives outside the local health system when perceived quality is low, effectively reducing their reported access. This interpretation is consistent with evidence from rural Nigeria showing that better-educated individuals are less reliant on dysfunctional primary healthcare services (Oluwadare *et. al*., 2023) and aligns with findings by Sou *et al*. (2023), who report lower engagement with formal health records among individuals with secondary or higher education.

Household size is positively associated with adequate healthcare access, a result that contrasts with conventional expectations that larger households face greater resource constraints. In rural Nigeria, this pattern may reflect adaptive coping strategies among large households, including reliance on low-cost, informal, or lower-quality healthcare providers that remain physically and financially accessible. While such choices increase the likelihood of reporting access, they may not correspond to improvements in service quality. This finding is inconsistent with Song *et. al*. (2025) but aligns with evidence reported by Anaeli *et. al*. (2025), highlighting the context-specific nature of household decision-making in constrained health systems.

Contrary to expectations, residence in rural northern Nigeria is associated with a higher probability of reporting adequate access and a lower probability of inadequate access to healthcare. This pattern likely reflects differential utilization of low-quality healthcare services rather than superior system performance in the North. In rural southern areas, relatively better-off households may opt out of low-quality local care in favor of private or higher-level services, thereby reducing their reported access within the local health system. By contrast, in the North—where a larger share of households are socioeconomically disadvantaged—available healthcare resources, including low-quality formal and informal services, may be more intensively utilized, increasing reported access despite persistent quality constraints (Ahuru *et. al*., 2021). Similar patterns have been observed in other low- and middle-income settings, where access indicators capture service reach rather than effective or high-quality care (Dong *et. al*., 2025).

Service delivery characteristics also play a critical role. Longer waiting times significantly reduce the probability of adequate access and increase the likelihood of inadequate access, underscoring waiting time as a major non-financial barrier to healthcare utilization. Extended waits impose substantial opportunity costs on rural households and may lead to delayed or foregone care, particularly for time-sensitive conditions (Findling *et. al*., 2020; Turnel *et. al*., 2022). This result is consistent with prior evidence from rural Nigeria (Nwokoro *et. al*., 2022) and from high-income settings, where disadvantaged populations face longer waiting times even within the same facilities (Anselmi *et. al*., 2017).

Conversely, longer facility operating hours are associated with a higher probability of adequate access and a lower likelihood of inadequate access. Extended hours likely improve temporal availability, allowing households to seek care without sacrificing work or household responsibilities, thereby facilitating timely treatment and potentially reducing avoidable complications. This finding is consistent with studies showing that flexible or extended clinic hours improve access to primary care and reduce reliance on emergency services (Johnson *et. al*., 2024).

Finally, perceived quality of healthcare is a strong predictor of access. Households that rate healthcare quality as good are significantly more likely to report adequate access and less likely to report inadequate access. This highlights the central role of perceived service quality in shaping healthcare-seeking behavior, beyond financial or geographic considerations. Similar associations between perceived quality and healthcare access have been documented in both low- and middle-income and high-income settings (Endalamaw *et. al*., 2023; Kukulka *et. al*., 2024).

## 5. Conclusion and Policy option

Given the multidimensional nature of healthcare access, selecting appropriate indicators that capture its key dimensions is essential for addressing persistent barriers to universal health coverage. This study developed and validated a psychometric measure of healthcare access and applied it to primary data from 624 rural households in Nigeria to identify factors associated with different levels of access. By combining a multidimensional access scale with robust empirical methods, the study provides policy-relevant evidence on structural and household-level constraints to healthcare access in rural settings.

The findings highlight several priority areas for policy action. First, the strong influence of consumption expenditure underscores the continued dominance of out-of-pocket financing, emphasizing the need to expand financial protection through wider health insurance coverage and reduced user fees. Second, persistent effects of service availability, waiting time, and operating hours point to critical supply-side constraints, calling for increased investment in rural health infrastructure, workforce capacity, and facility readiness, alongside operational reforms to improve service responsiveness. Third, the importance of perceived quality suggests that improving access requires more than physical proximity; strengthening service quality may enhance utilization and trust, particularly among more educated households. Finally, the reliance of larger and poorer households on low-cost or informal providers indicates that observed access often reflects coping strategies rather than effective care. Distinguishing nominal from quality-adjusted access and embedding multidimensional access measures into routine health surveys can strengthen evidence-based planning and support targeted efforts to reduce rural health inequities.

This study is subject to some limitations that warrant consideration when interpreting the findings. Fieldwork constraints arising from insecurity in parts of northern Nigeria limited full national representativeness; however, the persistence of a north–south socioeconomic structure and the application of appropriate sampling weights support the validity of population-level inference. In addition, the reliance on self-reported measures may introduce recall or measurement bias. While the absence of a universally accepted, context-comparable measure of healthcare access constrains external validation, the constructed multidimensional access scale demonstrated robustness across alternative estimation approaches. Finally, the indicators used to capture geographical accessibility were relatively limited, suggesting that future research would benefit from incorporating more objective spatial measures.

## Data Availability

The data used for this study are available are available on request from the corresponding author. The data are not available in the public domain due to privacy or ethical restrictions.

## Authors’ contributions

A.M, and K.K conceptualized the study. A.M and KK prepared and developed the proposal. K.K designed the sampling procedure. A.M performed the statistical analyses and prepared the first draft of the manuscript. A.M, and K.K reviewed the final version of the manuscript. All the authors have read and approved the final manuscript.

## Ethical approval and informed consent

Ethical approval for this study was obtained from the Nigeria National Health Research Ethics Committee (July 6, 2022) prior to data collection. All study procedures were conducted in accordance with relevant ethical guidelines and regulations. Participants were informed about the objectives of the study, and written informed consent was obtained from all participants before enrolment. Confidentiality and anonymity of the information provided were assured, and the data were used solely for research purposes.

## Appendix I: Original items on healthcare accessibility questionnaire

| S/N | Items | Dimension |
| --- | --- | --- |
| 1. | The facility has experts and skilled professionals | Availability |
| 2. | Has adequate equipment and facilities | Availability |
| 3. | Has sufficient staff including nurses and doctors | Availability |
| 4. | Has Sufficient bed space | Availability |
| 5. | Has easy and flexible mode of payment | Affordability |
| 6. | The facility provides affordable cost | Affordability |
| 7. | The facility provides easy accessibility | Geographical accessibility |
| 8. | Efficacy of drug and treatment is satisfactory | Acceptability |
| 9. | Effective for treating chronic diseases | Acceptability |
| 10. | Practitioners are usually cordial and friendly in handling patients | Acceptability |

## Appendix II: Standardized factor Loadings from Confirmatory Factor Analysis (CFA)

| Latent construct | Indicators (items) | Standard loadings |
| --- | --- | --- |
| Availability | Has adequate facility | 0.735 |
|  | Has sufficient staff including nurses & doctors | 0.845 |
|  | Has sufficient bed space | 0.82 |
| Affordability | Has easy and flexible mode of payment | 0.647 |
|  | The facility provides affordable cost | 0.731 |
| Geographical Accessibility | The facility provides easy accessibility | 0.652 |
| Acceptability | Efficacy of drug and treatment is satisfactory | 0.626 |
| Acceptability | Effective for treating chronic diseases | 0.535 |

Note: All coefficients are standardized estimates ( $\beta$ ) from the Structural Equation Model (SEM). Loadings below 0.40 are typically considered weak; all reported loadings are significant at $p < 0.05$ .

## APPENDIX III: Results of tests of instrument validity in Linear IV Model

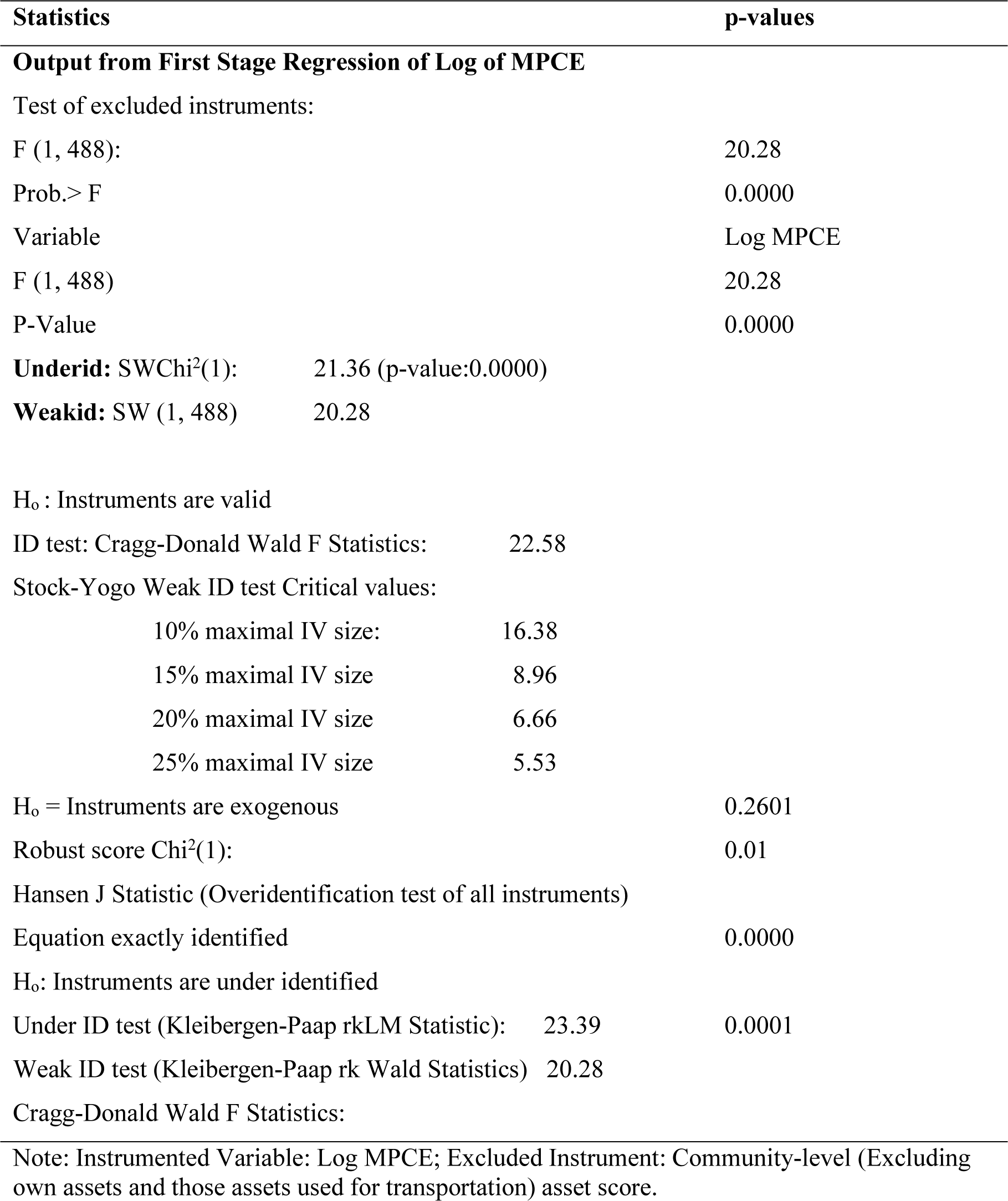

## References

Adam T, Ahmad S, Bigdeli M, Ghaffar A, Røttingen JA. 2023. Trends in health policy and systems research over the past decade: Still too narrow? Health Research Policy and Systems, 21: 34. 10.1186/s12961-023-00974-0

Adewuyi EO, Auta A, Adewuyi MI, Philip AA, Olutuase V, Zhao Y, Khanal V. 2024. Antenatal care utilisation and receipt of its components in Nigeria: Assessing disparities between rural and urban areas — a nationwide population-based study. PLoS ONE, 19 (7): e0307316. 10.1371/journal.pone.0307316

Afulani PA, Phillips B, Aborigo RA, Moyer CA. 2023..Person-centred quality of care and maternal health outcomes in low- and middle-income countries: A systematic review.BMJ Global Health, 8(3):e010890. 10.1136/bmjgh-2022-010890

Ahuru RR, Okungbowa OG, Iseghohi JO, Akpojubaro HE. 2021. Non-utilization of Primary Healthcare Centres for Skilled Pregnancy Care among Women in Rural Communities in Delta State, Southern Nigeria: Perspectives from Mothers, Fathers, and Healthcare Providers. Journal of International Women’s Studies, 22(9):

Andersen RM. 1995. Revisiting the Behavioral Model and Access to Medical Care: Does it Matter? Journal of Health and Social Behavior, 36(1): 1–10. 10.2307/2137284

Anaeli A, Kagaigai A, Luoga P, Nyamhanga T, Tungu M. 2025. The role of health insurance on healthcare utilisation among women in Tanzania: insights from Tanzania demographic and health survey. Arch Public Health 83(1):176. doi: 10.1186/s13690-025-01623-2.

Anselmi L, Meacock R, Kristensen SR, Doran T, Sutton M., 2017. Arrival by ambulance explains variation in mortality by time of admission: retrospective study of admissions to hospital following emergency department attendance in England. BMJ Qual. Saf. 26: 613–621. 10.1136/bmjqs-2016-005680

Daud KAM, Zulkarnaen KN, Rasdan IA, et. al. 2018. Validity and reliability of instrument to measure social media skills among small and medium entrepreneurs at Pengkalan Datu River. Int J Dev Sustain. 7(3): 1026–37

Dong B, Jiang MH, Zong J. 2025. Decomposition analysis of public health service utilization and health disparities among urban and rural older adult migrants in China. Front. Public Health 13:1591804. doi: 10.3389/fpubh.2025.1591804.

Edeh JN. 2022. Health insurance coverage and healthcare utilisation among households in Nigeria. BMC Health Services Research, 22: 1246.

Ensor T, Cooper S. 2004. Overcoming barriers to health service access: Influencing the demand side. Health Policy and Planning, 19(2): 69–79. 10.1093/heapol/czh009

Endalamaw, A., Khatri, R. B., Erku, D., Nigatu, F., Zewdie, A., Wolka, E., & Assefa, Y. 2023. Successes and challenges towards improving quality of primary health care services: a scoping review. BMC Health Services Research, 23: (893). 10.1186/s12913-

Filmer D, Pritchett LH. 2001. Estimating wealth effects without expenditure data—or tears: An application to educational enrollments in states of India. Demography, 38(1): 115–132. 10.1353/dem.2001.0003

Findling MG, Blendon RJ, Benson JM. 2020. Delayed care and unmet needs during the COVID-19 pandemic: Findings from a national survey in the United States. JAMA Health Forum, 1(12): e201463. 10.1001/jamahealthforum.2020.1463

Greene N. 2012. Econometric Analysis. New Jersey, USA: Prentice-Hall

Hair JF, Black WC, Babin BJ, et al. 2019. Multivariate data analysis (8thed.) Andover, Hampshire: Cengage Learning EMEA.

Hoseini-Esfidarjani SS, Negarandeh R, Delavar F. et al. 2021. Psychometric evaluation of the perceived access to health care questionnaire.BMC Health Services Research, 21(1): 638. 10.1186/s12913-

Hu LT, Bentler PM. 1999. Cutoff criteria for fit indexes in covariance structure analysis: Conventional criteria versus new alternatives. Structural Equation Modeling: A Multidisciplinary Journal, 6(1): 1–55. 10.1080/10705519909540118

Johnson C, Bourgoin D, Dupuis JB. 2024. Exploration of primary care models and timely access to care in New Brunswick (Canada). BMC Primary Care,25(366): 10.1186/s12875-024-02618-8

Knudsen AK, van den Berg R. 2024. The role of meso level characteristics of the health care system and socioeconomic factors on health care use – results of a scoping review. International Journal for Equity in Health, 23:149. 10.1186/s12939-024-02122-6.

Kukulka K, Benson JJ, Landon OJ, Makinde KW, Egginton B, Washington KT. A qualitative exploration of factors influencing healthcare utilization among rural Missourians: “We have to be bleeding, broken”. Health Place, 90: 103367. 10.1016/j.healthplace.2024.103367

Kumah E. 2022. The informal healthcare providers and universal health coverage in low- and middle-income countries. Globalization and Health, 18: 45. 10.1186/s12992-022-00839-z BioMed Central+1

Kruk ME, Gage AD, Arsenault C, Jordan K, Leslie HH., Roder-DeWan S, Pate M. (2018). High-quality health systems in the Sustainable Development Goals era: Time for a revolution. The Lancet Global Health, 6(11): e1196–e1252.

Kline RB. 2016. Principles and practice of structural equation modeling (4th ed.). New York, NY: The Guilford Press.

Levesque JF, Harris MF, Russell G. 2013. Patient-centred access to health care: Conceptualising access at the interface of health systems and populations. International Journal for Equity in Health, 12: 18. 10.1186/1475-9276-12-18

Macrotrend 2024. Nigeria rural population (% of total population): 1960–2024. Macrotrends LLC.https://www.macrotrends.net/countries/NGA/nigeria/rural-population

Martin CR, Emily S-MG 2013. A ‘good practice’guide for the reporting of design and analysis for psychometric evaluation. J Reprod Infant Psychol. 31(5):449–55.

Montgomery MR, Gragnolati M, Burke KA, Paredes E. 2000. Measuring living Standards with proxy variables. Demography, 37(2): 155–174. 10.2307/2648118

Mosha PE, Msengwa AS, Selemani M. 2025. Modeling determinants of accessibility for healthcare services in rural and urban areas of Dodoma, Tanzania. BMC Public Health, 25: 2920. 10.1186/s12889-025-22909-8.

Musau MM, Njogu, A, Maina A, Snow RW, Beňová L. et al. 2025. Methods for modelling composite indices of access to healthcare facilities: a systematic literature review. Population Health Metrics.

Mussa EC, Palermo T, Angeles G, Kibur M, Otchere F, Gavrilovic M, et al. Impact of community-based health insurance on health services utilisation among vulnerable households in Amhara region, Ethiopia. BMC Health Serv Res. 23(1):1–15.

National Population Commission NPC, ICF. 2023. Nigeria Demographic and Health Survey 2023: Key Indicators Report. Abuja, Nigeria, and Rockville, Maryland, USA: NPC and ICF.

Nwokoro UU, Ugwa OM, Ekenna AC, Obi IF, Onwuliri CD, Agunwa 2022. Determinants of primary healthcare services utilisation in an under-resourced rural community in Enugu State, Nigeria: A cross-sectional study. Pan African Medical Journal, 42, (209). 10.11604/pamj.2022.42.209.33317

Nwankwo ON, Ugwu CI, Nwankwo GI, Akpoke MA, Anyigor C, Obi-Nwankwo U, Spicer N. 2022. A qualitative inquiry of rural-urban inequalities in the distribution and retention of healthcare workers in southern Nigeria. PLoS ONE, 17(3): e0266159. 10.1371/journal.pone.0266159

Okunogbe A, Ogundeji YK., Adebayo EF. 2022. Out-of-pocket health expenditure and catastrophic spending in Nigeria. International Journal of Health Economics and Management, 22(4): 423–441. 10.1007/s10754-022-09329-5

Okoli C, Hajizadeh M, Rahman MM, Khanam R. 2020. Geographical and socioeconomic inequalities in the utilization of maternal healthcare services in Nigeria: 2003–2017. BMC Health Services Research, 20, Article 849. 10.1186/s12913-020-05700-w.

Olatunji SO, Ehebha EO Ifeanyi-Obi CC. 2013. Utilization of Western and Traditional Healthcare Services by Farm Families in Ukwa-East Local Government Area of Abia State. Journal of Agriculture and Social Research, 13 (2): 1–10.

Oluwadare A, Adepoju O. 2023. Awareness and utilization of primary healthcare services in a rural community in Nigeria. *Journal of Public Administration*, Policy and Governance Research (JPAPGR), 2(1): https://jpapgr.com/index.php/research/article/download/60/60/110

Onakalu PO, Oyinlola FF, Oluwatope OB, Agbeja I, Shittu IO, Ogbeye GB, Okorafor KA. 2025. Rural-urban differentials in women’s empowerment and experience of under-five mortality among mothers in Nigeria: a Multiple Indicator Survey analysis. BMC Public Health, 25, Article 2208. 10.1186/s12889-025-23412-w.

Onah MN, Govender V, McIntyre D. 2023. Assessing equity in health service utilization in Nigeria: Implications for universal health coverage.Health Policy and Planning, 38 (2): 214–226. 10.1093/heapol/czac092

Onwujekwe O, Mbachu C, Onyebueke V, Ogbozor P, Arize I, Okeke C, Ezenwaka U, ; Ensor T. 2022. Stakeholders’ perspectives and willingness to institutionalize linkages between the formal health system and informal healthcare providers in urban slums in southeast Nigeria. BMC Health Services Research, 22: 583. 10.1186/s12913-022-08005-2

Palmer N. 2007. Access and equity: evidence on the extent to which health services address the needs of the poor. In: Bennett S, Gibson L and Mills A, editors, Health, economic development and household poverty, London: Routledge, 61–73.

Peters DH, Garg A, Bloom G, Walker DG, Brieger WR, Rahman MH. 2008. Poverty and access to healthcare in developing countries. Ann. N Y Acad. Sci. 1136:161–171.

Penchansky R and Thomas JW. 1981. The concept of access: definition and relationship to consumer satisfaction. Medical Care 19: 127–140.

Pengpid S, Peltzer K. 2021. The prevalence and correlates of suicidal behaviour among adolescents in Trinidad and Tobago. Journal of Psychology in Africa, 31(4):424–429. 10.1080/14330237.2021.1928926

Porgo TV, Amissah RQ, Bachongy K, Sorgho G, Kpegli YT.2024. Harnessing the power of the private sector for universal health coverage in Sub-Saharan Africa. World Bank Blogs. https://blogs.worldbank.org.

Population Reference Bureau. 2024. Universal Health Coverage (UHC) Service Coverage Index by country: Nigeria, Ghana, Senegal, and Côte d’Ivoire. Population Reference Bureau. Retrieved from https://www.prb.org/international/indicator/uhcindex/snapshot

Suo Z, Shao L, Lang Y. 2023. A study on the factors influencing the utilization of public health services by China’s migrant population based on the Shapley value method. BMC Public Health 23: 2328 10.1186/s12889-023-17193-3.

Song Z. 2025. Paying Primary Care More — Will It Work This Time? JAMA. Advance online publication. 10.1001/jama.2025.16467

Tanou M, Kishida T, Kamiya Y. 2021. The effects of geographical accessibility to health facilities on antenatal care and delivery services utilization in Benin: A cross-sectional study. Reproductive Health, 18 (205). 10.1186/s12978-021-01249-x

Turner AJ, Francetic I, Watkinson R, Gillibrand S, Sutton M. 2022.Socioeconomic inequality in access to timely and appropriate care in emergency departments. Journal of Health Economics, 85: 102668.

World Bank. 2024. Nigeria country overview — population and urbanization. World Bank Data. Retrieved February 2026, from https://www.data.worldbank.org/country/nigeria

Wossen T, Alene AD, Abdoulaye T. 2018. Poverty reduction effects of technology Adoption: Evidence from Nigeria. Proceedings of the International Association of Agricultural Economists Conference, July 28–August 2, Vancouver, British Columbia. 10.22004/ag.econ.276980

Wooldridge JM. 2015. Introductory econometrics: A modern approach (6th ed.). Mason, OH: South-Western, Cengage Learning

Whitehead M, Dahlgren G, Evans T. 1997. Equity and health sector reforms: Can low-income countries escape the medical poverty trap? Lancet, 358(9284): 833–836.

World Health Organization. 2025. Universal health coverage and access to health services: Definitions and measurement. World Health Organization.

World Health Organization & World Bank. 2023. Billions left behind on the path to universal health coverage (UHC Global Monitoring Report). World Health Organization.

World Health Organization, UNICEF, World Bank Group. 2023. *Trends in maternal mortality:* 2000 to 2023 (WHO, UNICEF, World Bank Group report). World Health Organization.

Weitensfelder L, Moshammer H, Ataniyazova O. 2024. Energy consumption, energy distribution, and clean energy use together affect life expectancy. Sustainability, 16:678.

World Health Organization. 2025. *Tracking Universal Health Coverage*: 2025 Global Monitoring Report. World Health Organization & World Bank Group.

